# Hepatitis C prevalence and key population size estimate updates in San Francisco: 2015 to 2019

**DOI:** 10.1101/2021.10.24.21265448

**Authors:** Shelley N. Facente, Rachel Grinstein, Roberta Bruhn, Zhanna Kaidarova, Erin Wilson, Jennifer Hecht, Katie Burk, Eduard Grebe, Meghan D. Morris, on behalf of End Hep C SF

**Author notes:** Corresponding author (S.N.F.).

## Abstract

**BACKGROUND:** With the introduction of direct-acting antivirals to treat and cure hepatitis C virus (HCV) infection, HCV elimination is achievable. In 2017, San Francisco’s HCV elimination initiative, *End Hep C SF*, generated an estimate of city-wide HCV prevalence in 2015, but only incorporated limited information about population HCV treatment. Using additional data and updated methods, we aimed to update the 2015 estimate to 2019 and provide a more accurate estimate of the number of people with untreated, active HCV infection overall and in key subgroups – people who inject drugs (PWID), men who have sex with men (MSM), and low socioeconomic status transgender women (low SES TW).

**METHODS:** Our estimates are based on triangulation of data from blood bank testing records, cross-sectional and longitudinal observational studies, and published literature. We calculated subpopulation estimates based on biological sex, age and/or HCV risk group. When multiple sources of data were available for subpopulation estimates, we calculated an average using inverse variance weighting. Plausible ranges (PRs) were conservatively estimated to convey uncertainty.

**RESULTS:** The total number of people estimated to have anti-HCV antibodies in San Francisco in 2019 was 22,585 (PR:12,014–44,152), with a citywide seroprevalence of 2.6% (PR:1.4%–5.0%) – similar to the 2015 estimate of 21,758 (PR:10,274–42,067). Of all people with evidence of past or present infection, an estimated 11,582 (PR:4,864–35,094) still had untreated, active HCV infection, representing 51.3% (PR:40.5%–79.5%) of all people with anti-HCV antibodies, and 1.3% (PR:0.6%–4.0%) of all San Franciscans. PWID comprised an estimated 2.8% of the total population of San Francisco, yet 73.1% of people with anti-HCV antibodies and 90.4% (n=10,468, PR:4,690–17,628) of untreated, active HCV infections were among PWID. MSM comprised 7.8% of the total population, yet 11.7% of people with anti-HCV antibodies and 1.0% (n=119, PR:0–423) of those with untreated active infections. Low SES TW comprised an estimated 0.1% of the total population, yet 1.4% of people with HCV antibodies and 1.6% (n=183, PR:130–252) of people with untreated active infections.

**CONCLUSIONS:** Despite the above-average number (2.6%) of people with anti-HCV antibodies, we estimate that only 1.3% (PR:0.6%–4.0%) of all San Francisco residents have untreated, active HCV infection – likely a reflection of San Francisco’s robust efforts to diagnose infection among high-risk groups and initiate curative treatment with as many people as possible. While plausible ranges of infections are wide, these findings indicate that while the overall number of people with anti-HCV antibodies may have increased slightly, the number of people with active HCV infection may have decreased slightly since 2015. This estimate improves upon the 2015 calculations by directly estimating the impact of curative treatment citywide and in subgroups. However, more research is needed to better understand the burden of HCV disease among other subgroups at high risk, such as Blacks/African Americans, people with a history of injection drug use (but not injecting drugs in the last 12 months), people who are currently or formerly incarcerated, and people who are currently or formerly unhoused.

## Introduction

Since the introduction in late 2013 of highly effective direct-active antivirals (DAAs) for treatment and cure of hepatitis C virus (HCV) infection [1], elimination of HCV is now an achievable goal, and HCV elimination initiatives have been launched at the international [2], U.S. national [3], and local levels [4,5]. Despite these efforts, the global burden of HCV infection remains massive, with approximately 1% of the world population chronically infected with HCV, an estimated 71 million people [6] – around twice the number of people living with HIV, which cannot be cured [7]. In the U.S., rates of acute HCV infection have been increasing since 2009, especially in young adults. In 2018, there was a bimodal distribution by age of newly reported chronic HCV cases reported to the National Notifiable Diseases Surveillance System, with the greatest number of newly reported cases among persons aged 20-39 and 50-69 [8].

San Francisco announced its own HCV elimination initiative in 2016, known as *End Hep C SF* [5]. One early task of the Research and Surveillance workgroup of *End Hep C SF* was to calculate the first-ever estimate of the number of people living with HCV in the city at the start of the initiative, with particular emphasis on the burden of disease for people who inject drugs (PWID), men who have sex with men (MSM), and transgender women (TW), and the remaining population disaggregated by age and sex [9]. However, at that time there was little available data related to HCV treatment for people in San Francisco, and as such the estimate of people with active infection was unable to incorporate the impact of received treatment within subgroups, and instead relied upon a total overall sum of known treatment initiations citywide, based on summarized medical record data from a portion of the city’s medical systems.

In 2020 – approximately six years after DAAs became available in San Francisco and five years after the launch of *End Hep C SF* – we initiated an update of the 2015 estimate of HCV prevalence for 2019 that incorporates expanded treatment data from medical records and observational research studies to estimate the remaining population of untreated, active HCV infection, overall and within key subgroups.

## Methods

As with the 2015 estimate, we used a triangulation approach for this analysis [10,11], synthesizing multiple overlapping data sources to produce reliable estimates of the number of people in San Francisco with active HCV infection, accounting for those with (1) acute infection (i.e., recently infected people who may still spontaneously clear infection [12] or may become chronically infected), and (2) chronic infection, in a variety of population strata, after excluding an estimated number of people who were chronically infected but have been treated and cured in each stratum. Sources of local HCV-related data included blood bank infectious disease testing records, cross-sectional and longitudinal observational studies, and published literature gathered through a systematic review. As in 2015, plausible ranges (PRs) for all estimates were derived from 95% confidence intervals (CIs) of population size and prevalence estimates (i.e., the lower bound of the PR is the lower bound of the 95% CI for the population size of a subgroup multiplied by the lower bound of the 95% CI for the prevalence estimate for that subgroup and vice versa), resulting in conservative (intentionally wide) PRs. As all data used were de-identified or already aggregated and no human participants were involved in this study, we did not seek approval from an ethics board to carry out this work.

### Literature Review

For this 2019 update, we used the identical search terms as in 2015, but changed the publication date range to January 2017 through January 2021 in MEDLINE and Science Citation Index Expanded, to append new articles to the prior search (Table S1). These searches resulted in 34 and 245 new articles found, respectively. Each abstract was then reviewed by S.N.F, discarding those that were not specific to San Francisco or otherwise did not provide estimates relevant for the data triangulation employed in this analysis. Following de-duplication, 11 unique abstracts were retained for incorporation into the estimate (Table S2); however, of those 11 only four [13-16] had estimates that were utilized in the data triangulation, as the other seven were new publications from original datasets available for direct analysis (i.e., data from the UFO study, National HIV Behavioral Surveillance (NHBS) datasets from the 2018 PWID wave and 2017 MSM wave, and datasets from the Transfemales Empowered to Advance Community Health 3 (TEACH3) study and the trans-focused wave of NHBS in 2019, also known as TEACH4, both of which focus on TW with low socioeconomic status, or SES). Four articles included in the 2015 estimate were also incorporated into this 2019 update [17-20], as they were published within the past six years and provided unique data points for the analysis.

### Stratification of the San Francisco Population

The 2019 American Community Survey (ACS) informed the overall and general population size estimates (PSE) [21]. Like with the 2015 estimate, we separately estimated population sizes for key subgroups bearing high HCV burden, including PWID, MSM, and TW with low SES. Remaining residents were then considered part of a ‘general population,’ which included children; adults who are at a low risk of living with HCV; and adults who have a history of drug injection (but have not injected drugs in the past 12 months), are currently or formerly homeless, and/or are currently or formerly incarcerated. To address uneven distribution of infection by age and sex we stratified the general population as follows: children 14 and under (both sexes), and males and females separately for ages 15-54, 55-74 (which in 2019 included people born 1945-1965, known as the “birth cohort” of baby boomers), and 75 and older.

### Population Size Estimates

#### PWID

As no updated information was available to inform PWID population size estimates (PSEs) since the 2015 analysis and there were no trends identified that would dramatically affect mortality or in- or out-migration of PWID from 2015 to 2019, the same methods used in the 2015 analysis were retained here [9]. We again calculated a weighted average of the individual estimates available in Chen *et al*. 2016 [17], using inverse variance weighting and assuming the CIs of those estimates had a normal distribution. Like in the 2015 analysis, we excluded two estimates that appeared to be considerably biased and would have skewed the weighted average [9].

#### MSM

We similarly calculated an inverse variance weighted average of the MSM PSE using estimates from the literature, both the two papers used in 2015’s estimate [19,20] and incorporating a new article estimating 19.4% more MSM living in San Francisco in 2017, compared to earlier estimates from 2014 [13]. For a sub-analysis, we assumed that 18.8% of MSM in San Francisco were living with HIV [22]. While there is some overlap between the PWID and MSM strata, work specific to San Francisco has found distinct differences and limited population mixing between the group of MSM who inject drugs and generally appear in MSM-focused studies, and the group of PWID who are MSM and generally appear in PWID-focused studies (Artenie *et al*., under review), indicating that double-counting of individuals in this analysis is likely to be minimal.

#### Low SES TW

Like with the 2015 estimate, the weighted average for the PSE of low SES TW was mostly informed by multiplier estimates included in Wesson *et al*. [15], excluding the successive sampling estimate for consistency with our PWID PSE method. For 2019 we also added a new literature-based estimate for the proportion of the adult population that is TW from Crissman *et al*. [16] applied to the ACS data for San Francisco adults, then multiplied by the proportion of the TW population deemed to be of ‘low socioeconomic status’ in San Francisco [14], to make this estimate comparable to the subgroup of TW addressed in the Wesson analysis.

#### General Population

We generated point estimates and 95% confidence intervals in 5-year age categories for the 2019 ACS in San Francisco by sex, then deducted the estimated number of people who were PWID, MSM, or TW in each sex and age stratum using the age and sex distribution of participants in the NHBS 2017 MSM and 2018 PWID and 2019 TW San Francisco waves. We then aggregated the PSEs for the non-key population adult age/sex strata into three age categories for each sex, to allow for differentiation of those in the birth cohort.

### Anti-HCV Seroprevalence Estimates

#### PWID

We calculated a weighted average of three PWID HCV seroprevalence estimates using inverse variance weighting (fixed effects model); two estimates were calculated directly from HCV serological testing in the 2018 wave of NHBS and the UFO Study from 2016-2018, which enrolls PWID under age 30. One estimate was from a San Francisco methadone clinic-based study published in 2014[18]. Like in 2015, the fixed effects model was chosen since the studies were deemed to be imperfect measurements of the same underlying seroprevalence.

#### MSM

We similarly calculated an inverse variance weighted average for MSM seroprevalence, using three estimates: (1) direct calculation of the proportion of people who self-reported as HCV seropositive in the 2017 MSM wave of NHBS, (2) calculation of the proportion of people with HCV positive serologies listed in the electronic medical record from 2017-2018 for the Strut sexual health clinic run by the San Francisco AIDS Foundation (either from self-report or in-house testing), and (3) calculation of the proportion of anti-HCV positive results through community-based organization or clinic-focused testing funded by the San Francisco Department of Public Health (SFDPH), excluding testing at Strut or in the county jails. We also estimated the number of MSM in San Francisco who were co-infected with HCV and HIV, by taking the number of chronically infected MSM (estimated as described below) and estimating the number of MSM living with HIV who likely cleared the hepatitis C virus spontaneously, using literature-based estimates indicating that fewer MSM with HIV clear HCV spontaneously than those who are HIV-negative [23,24].

#### Low SES TW

For low SES TW, we directly calculated a weighted average of the proportion of participants in TEACH3 and TEACH4 who tested positive for anti-HCV antibodies during the studies, disregarding TEACH4 data from participants who were already included in the TEACH3 estimate, as an indicator of TEACH3 participation was available in the TEACH4 dataset.

#### General Population

Data for first-time allogeneic blood donors at Vitalant from 2010-2019 who were San Francisco residents at the time of donation (n=29,387) informed the seroprevalence estimate for non-PWID, MSM, or TW adults over 14 years of age. Donors’ ages were standardized to age in 2019 to match the 2019 ACS, and seroprevalence estimates were calculated for 5-year age bins, stratified by both sex and race. These seroprevalences were then collapsed into sex-stratified age categories for donors age 15-54, 55-74, and 75+, weighting the collapsed estimates by race using the racial proportions of HCV positive results newly reported to the San Francisco Department of Health from 2018-2019 for public health surveillance purposes [25], to better account for known racial disparities in HCV prevalence in San Francisco. To account for potential bias from the ‘healthy donor effect’ [26] and selection bias related to blood donor eligibility criteria (which exclude MSM, and others) we adjusted the directly calculated prevalence by an ‘inflation factor’. To estimate the inflation factors for males and females, we compared HCV seroprevalence in first-time blood donors reported by the national TTIMS study [27] to the 2018 National Health and Nutrition Examination Survey (NHANES) estimate calculated from participants age 16 years and older [28]. We removed respondents who reported ever injecting drugs from the NHANES data before estimating prevalence, though at the time of this analysis the relevant survey file allowing exclusion of MSM was not yet available. We used population weights and clustering was taken into account when calculating prevalence point estimates and standard errors from NHANES data. The ratio between TTIMS seropositivity and NHANES seropositivity was 7.3 for females and 14.3 for males. We set the plausible bounds of this ratio to be 50% in either direction, and used the resulting point estimate and intervals (7.3, PR 3.6-10.9 for females and 14.3, PR 7.2 – 21.5 for males) as the inflation factors by which Vitalant seroprevalences were multiplied for each of the sex/age categories in the final analysis.

#### Children

No known HCV seroprevalence data exist for San Francisco children under age 15; thus we used the same methods described in the 2015 estimate [9] to extrapolate the number of children likely seropositive for anti-HCV antibodies from the San Francisco-specific number of PWID women of childbearing age, the number of non-PWID women of childbearing age, the estimated prevalence of chronic HCV infection for each of these groups, the number of live births in the past 15 years in San Francisco [29], and a weighted average of the risk of vertical transmission of HCV for HIV-negative women in the US (0.040, 95% CI: 0.01 – 0.071) [30,31]. Rates of vertical transmission appear to differ by HIV status [32]; however, the number of HIV-infected women in San Francisco is extremely small [33].

### Estimates of Chronic HCV Infections

#### PWID

We calculated the estimated prevalence of chronic infection by taking an inverse variance weighted average of the proportion of positive results from RNA testing for PWID in both the UFO study from 2016-2018 and the 2018 NHBS wave.

#### MSM

The only available estimate of chronic HCV infection specifically for MSM was available through a dataset of SFDPH-funded community-based and clinic testing in 2019 (excluding jail-based testing), so the proportion of RNA tests in MSM that were positive in this dataset was used directly. While some individuals may have had repeated testing, we believe the number of repeat testers to be very small in this dataset given the time restriction to one calendar year, and not enough to substantially bias the proportion used. The estimated number of chronically infected MSM was then multiplied by the proportion of MSM in the 2017 NHBS wave who reported being co-infected with both HIV and HCV to estimate the prevalence of chronic HCV infection among MSM living with HIV.

#### Low SES TW

Like with seroprevalence, we directly calculated a weighted average of the proportions of TW in the TEACH3 and TEACH4 studies who tested positive for HCV RNA.

#### General Population

The same methods for calculating seroprevalence from Vitalant blood donor data were used for estimating a race-weighted chronic prevalence for the same age/sex combinations, this time counting donations where donors were both anti-HCV antibody and RNA positive, assuming the vast majority of those were indeed chronically infected and unlikely to clear the virus spontaneously.

#### Children

Like in 2015, we used an estimate of chronic HCV prevalence for children older than 18 months from a meta-analysis of mother-to-child transmission, published in 2014 [32] to estimate the proportion of children with anti-HCV antibodies who likely progressed to chronic infection. Chronic infection was defined as the child testing positive for HCV RNA at older than 18 months (when maternal antibodies have waned and spontaneous clearance would typically have already occurred. As we had no direct estimate of seroprevalence among children or of the proportion of perinatally-infected children who spontaneously clear the infection, the chronic prevalence was used as a conservative estimate of seroprevalence.

### Estimates of Acute HCV Infections

#### PWID

Because of the longitudinal nature of the UFO study with regular intervals of HCV RNA testing, we were able to directly calculate the HCV incidence rate for participants in the cohort between 2017-2019 (10 cases over 57,678 person-days of observation). We then used that information to directly estimate the prevalence of acute HCV that would have accumulated in the UFO study population over a 6-month period (i.e., the average length of time between initial infection and being considered chronically infected) and used that figure as a proxy for acute HCV prevalence in all PWID.

#### MSM

No comparable dataset was available for calculating the HCV incidence rate among MSM; therefore, as in the 2015 analysis we used HCV incidence estimates for MSM reported by Yaphe *et al*. [34], assuming time-invariant incidence and a mean RNA positive/antibody negative window of 60 days. HCV window period ranges were then used as the best available proxy to estimate the credible range of MSM acute prevalence.

#### Low SES TW

For TW, no data were found for acute infection prevalence or incidence of HCV infection. Therefore, like as for the 2015 estimate, TW acute prevalence is approximated as having the same ratio as TW to PWID seroprevalence. We again set the acute prevalence plausible range bounds to the point estimate for MSM acute prevalence and PWID acute prevalence, respectively, reflecting the general belief that acute prevalence for TW is higher than MSM but lower than PWID in San Francisco.

#### General Population

The same methods were used for estimating a race-weighted acute infection prevalence from Vitalant blood donor data for the same age/sex combinations, this time counting donations where donors were both RNA positive but anti-HCV antibody negative, again as the best available proxy for acute infection.

#### Children

Like for the 2015 analysis, we assumed no children were acutely infected with HCV at the point-in-time relevant to this analysis, given the rarity of HCV infection among children in San Francisco and the transient nature of acute infection.

### Estimates of People Treated and Cured

#### PWID

We calculated the proportion of people in both the most recent wave of the UFO study and 2018 PWID wave of NHBS who self-reported having completed HCV treatment, and took a weighted average of those proportions by age (age 29 and under vs. age 30 and over), recognizing the treatment proportions were considerably different between the UFO study (which only enrolls participants under age 30) and the NHBS wave overall. To determine the number treated from the time of the survey through the end of 2019, we needed to estimate a treatment rate for PWID in San Francisco from 2015 through 2019, since that rate is assumed to *not* be constant. Because no data were available for these estimates in San Francisco, multiple authors (S.N.F., M.M., E.G.) used estimates for treatment for the State of California in 2015-2017 available through mappinghepc.com, and worked together using subject matter expertise to design a plausible curve for San Francisco (see Figure 1). While the estimates of the absolute proportions treated in Figure 1 may not match the reality of treatment coverage in PWID each year, it is only important that the shape of the curve approximately reflects relative treatment rates at different times. We then used that curve to estimate the proportion of people treated between the survey and end of 2019, given the proportion of participants who reported treatment between 2015 and the date of the survey. That was used as the proportion of people counted as chronically infected in each stratum who were then considered subsequently cured and no longer having active infection as of the end of 2019.

**Figure 1.**
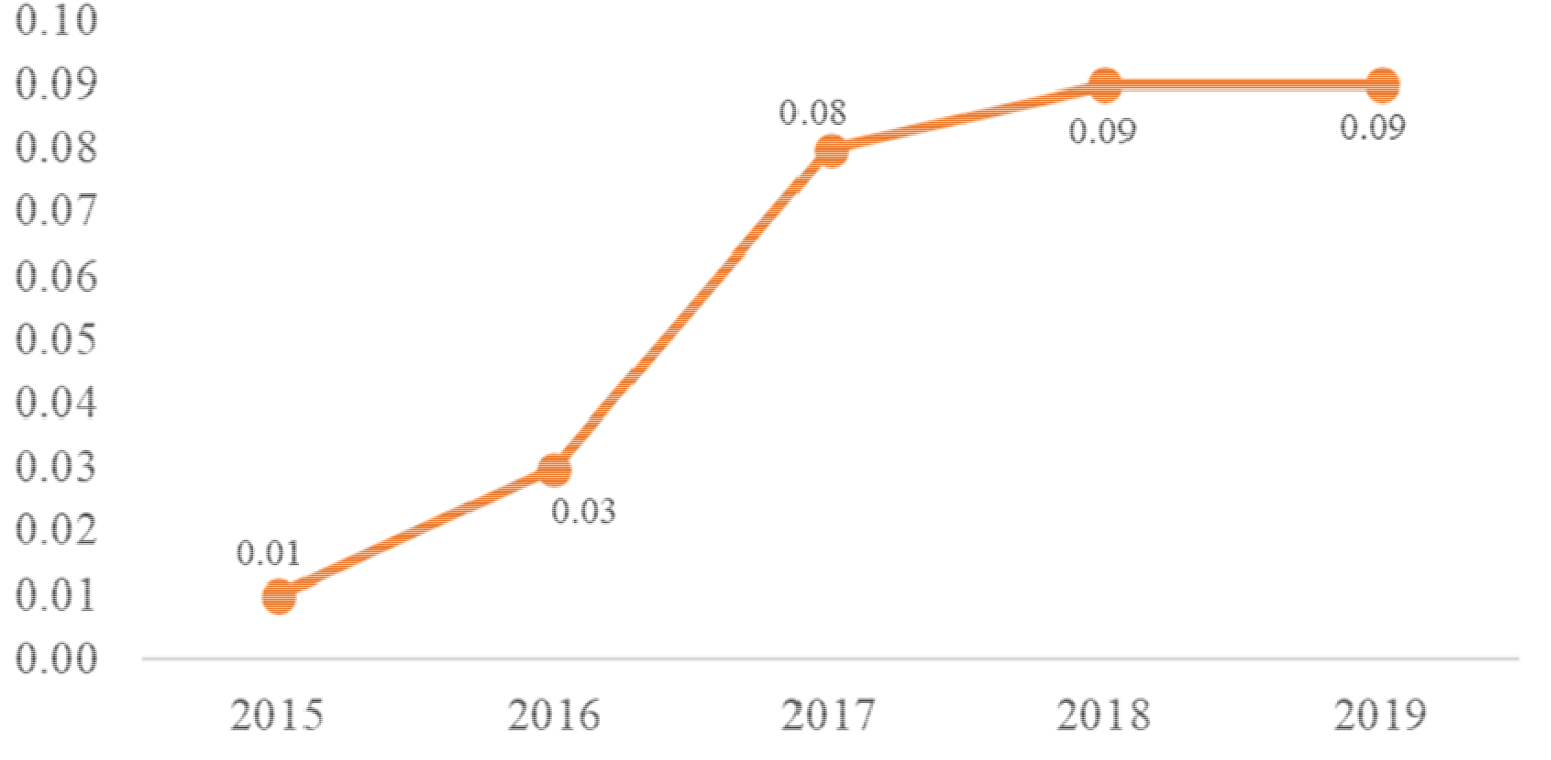
Estimated proportion of people who inject drugs, men who have sex with men, and trans women estimated to have been treated and cured of hepatitis C virus infection each year in San Francisco, 2015-2019.

#### MSM

We followed methods nearly identical to those described for PWID, except the proportion of people having completed treatment from 2015 through the survey dates in 2018 was an inverse variance weighted average of the proportions of participants in the 2017 NHBS wave who self-reported having completed HCV treatment, and the proportions of male patients of the Strut sexual health clinic who either received HCV treatment through the clinic or self-reported having been treated between 2017 and 2018. The estimated treatment curve used for PWID (Figure 1) was assumed to be the same for MSM.

#### Low SES TW

The same methods and same estimated treatment curve were used for low SES TW, using an inverse variance weighted average of self-reported treatment data from TEACH 3 and non-duplicate participants in TEACH 4.

#### General Population

The same methods as described above were used for estimating the prevalence of people with cleared or cured infections in the Vitalant blood donor population for the same age/sex combinations, this time counting donations where donors were anti-HCV positive but RNA negative. A weighted average of the proportion of people infected with HCV who were expected to spontaneously clear infection was then estimated for each of the age/sex categories from relevant literature where clearance estimates had been disaggregated by age and sex [35,36]. The proportion likely cleared in each category was subtracted from the total proportion of people with cleared or cured infections in the Vitalant dataset, and the remaining number of people were assumed to have been treated and cured.

#### Children

As DAAs were not FDA-approved for children under 18 years of age until March 2020 [37], we assumed no children had been treated for HCV in 2019, for purposes of this analysis.

## Results

Overall, the total number of people estimated to have anti-HCV antibodies in San Francisco in 2019 was 22,585 (plausible range [PR]: 12,014 – 44,152), with a citywide seroprevalence of 2.6% (PR: 1.4% – 5.0%). This represents a slight increase over the estimate of seroprevalence in 2015, though well within the wide PR bounds of the prior estimate (Table 1). Of those, 5,982 (PR: 3,238 – 11,430) were baby boomers, which represents an estimated seroprevalence of 3.2%. In terms of gender, 16,467 (PR: 8,741 – 31,148) people with anti-HCV antibodies were men, and 5,798 (PR: 3,019 – 12,632) were women (see Table 2), representing an estimated seroprevalence of 3.7% and 1.3%, respectively. Of all people with evidence of past or present infection (i.e., anti-HCV antibodies) in 2019, an estimated 11,582 (PR: 4,864 – 35,094) still had untreated, active HCV infection, representing 51.3% (PR: 40.5% - 79.5%) of all people with anti-HCV antibodies, and 1.3% (PR: 0.6% - 4.0%) of all San Francisco residents.

**Table 1.**
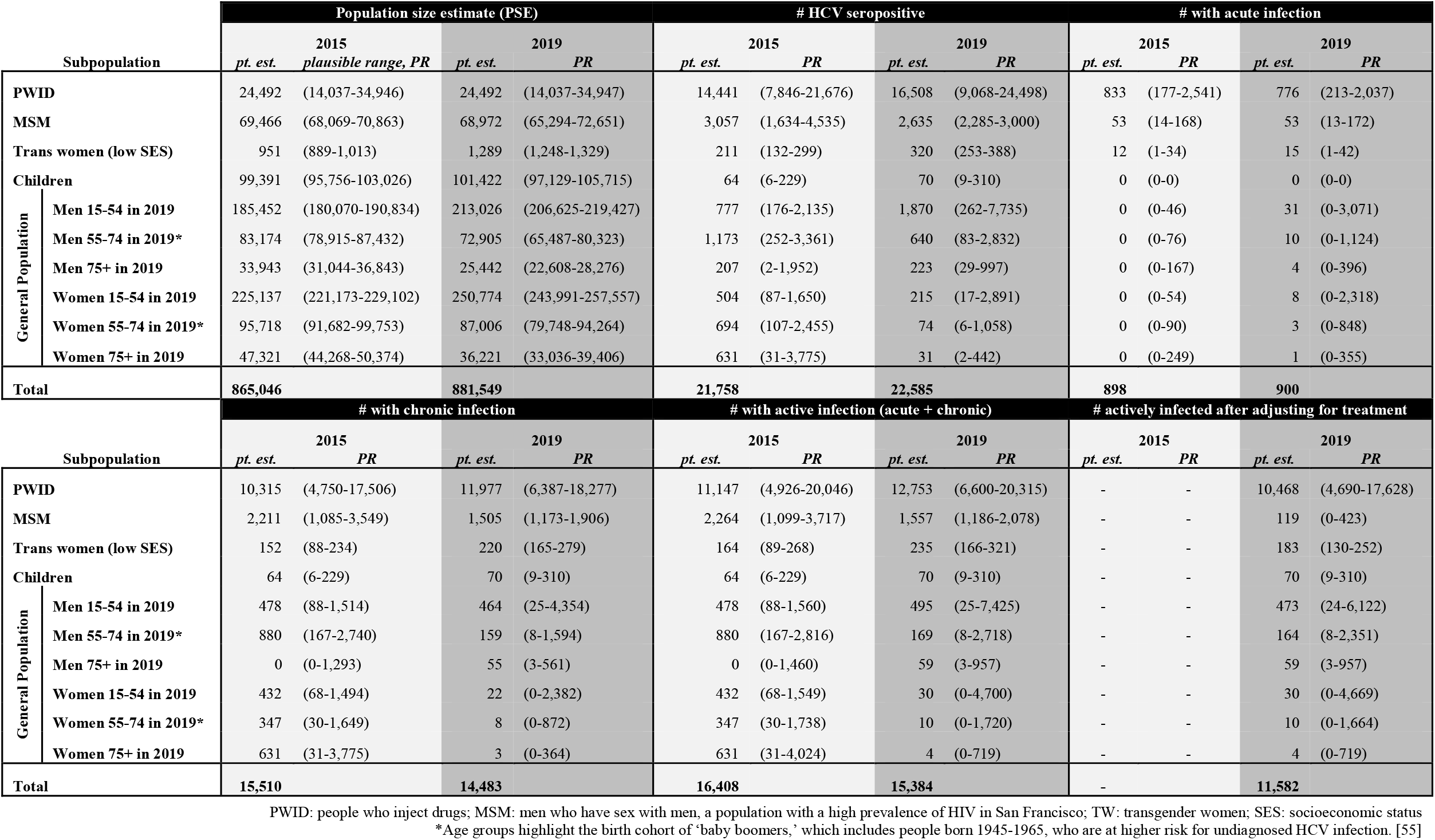
Estimated Population Size and Number of People with anti-HIV Seropositivity, Acute Infection, Chronic Infection, and Active Infection after Adjusting for Treatment, 2019.

| Subpopulation |  | Population size estimate (PSE) |  |  |  | # HCV seropositive |  |  |  | # with acute infection |  |  |  |
| --- | --- | --- | --- | --- | --- | --- | --- | --- | --- | --- | --- | --- | --- |
|  |  | 2015 |  | 2019 |  | 2015 |  | 2019 |  | 2015 |  | 2019 |  |
|  |  | *pt. est.* | *plausible range, PR* | *pt. est.* | *PR* | *pt. est.* | *PR* | *pt. est.* | *PR* | *pt. est.* | *PR* | *pt. est.* | *PR* |
| PWID |  | 24,492 | (14,037-34,946) | 24,492 | (14,037-34,947) | 14,441 | (7,846-21,676) | 16,508 | (9,068-24,498) | 833 | (177-2,541) | 776 | (213-2,037) |
| MSM |  | 69,466 | (68,069-70,863) | 68,972 | (65,294-72,651) | 3,057 | (1,634-4,535) | 2,635 | (2,285-3,000) | 53 | (14-168) | 53 | (13-172) |
| Trans women (low SES) |  | 951 | (889-1,013) | 1,289 | (1,248-1,329) | 211 | (132-299) | 320 | (253-388) | 12 | (1-34) | 15 | (1-42) |
| Children |  | 99,391 | (95,756-103,026) | 101,422 | (97,129-105,715) | 64 | (6-229) | 70 | (9-310) | 0 | (0-0) | 0 | (0-0) |
| General Population | Men 15-54 in 2019 | 185,452 | (180,070-190,834) | 213,026 | (206,625-219,427) | 777 | (176-2,135) | 1,870 | (262-7,735) | 0 | (0-46) | 31 | (0-3,071) |
|  | Men 55-74 in 2019* | 83,174 | (78,915-87,432) | 72,905 | (65,487-80,323) | 1,173 | (252-3,361) | 640 | (83-2,832) | 0 | (0-76) | 10 | (0-1,124) |
|  | Men 75+ in 2019 | 33,943 | (31,044-36,843) | 25,442 | (22,608-28,276) | 207 | (2-1,952) | 223 | (29-997) | 0 | (0-167) | 4 | (0-396) |
|  | Women 15-54 in 2019 | 225,137 | (221,173-229,102) | 250,774 | (243,991-257,557) | 504 | (87-1,650) | 215 | (17-2,891) | 0 | (0-54) | 8 | (0-2,318) |
|  | Women 55-74 in 2019* | 95,718 | (91,682-99,753) | 87,006 | (79,748-94,264) | 694 | (107-2,455) | 74 | (6-1,058) | 0 | (0-90) | 3 | (0-848) |
|  | Women 75+ in 2019 | 47,321 | (44,268-50,374) | 36,221 | (33,036-39,406) | 631 | (31-3,775) | 31 | (2-442) | 0 | (0-249) | 1 | (0-355) |
| Total |  | 865,046 |  | 881,549 |  | 21,758 |  | 22,585 |  | 898 |  | 900 |  |
| Subpopulation |  | # with chronic infection |  |  |  | # with active infection (acute + chronic) |  |  |  | # actively infected after adjusting for treatment |  |  |  |
|  |  | 2015 |  | 2019 |  | 2015 |  | 2019 |  | 2015 |  | 2019 |  |
|  |  | *pt. est.* | *PR* | *pt. est.* | *PR* | *pt. est.* | *PR* | *pt. est.* | *PR* | *pt. est.* | *PR* | *pt. est.* | *PR* |
| PWID |  | 10,315 | (4,750-17,506) | 11,977 | (6,387-18,277) | 11,147 | (4,926-20,046) | 12,753 | (6,600-20,315) | - | - | 10,468 | (4,690-17,628) |
| MSM |  | 2,211 | (1,085-3,549) | 1,505 | (1,173-1,906) | 2,264 | (1,099-3,717) | 1,557 | (1,186-2,078) | - | - | 119 | (0-423) |
| Trans women (low SES) |  | 152 | (88-234) | 220 | (165-279) | 164 | (89-268) | 235 | (166-321) | - | - | 183 | (130-252) |
| Children |  | 64 | (6-229) | 70 | (9-310) | 64 | (6-229) | 70 | (9-310) | - | - | 70 | (9-310) |
| General Population | Men 15-54 in 2019 | 478 | (88-1,514) | 464 | (25-4,354) | 478 | (88-1,560) | 495 | (25-7,425) | - | - | 473 | (24-6,122) |
|  | Men 55-74 in 2019* | 880 | (167-2,740) | 159 | (8-1,594) | 880 | (167-2,816) | 169 | (8-2,718) | - | - | 164 | (8-2,351) |
|  | Men 75+ in 2019 | 0 | (0-1,293) | 55 | (3-561) | 0 | (0-1,460) | 59 | (3-957) | - | - | 59 | (3-957) |
|  | Women 15-54 in 2019 | 432 | (68-1,494) | 22 | (0-2,382) | 432 | (68-1,549) | 30 | (0-4,700) | - | - | 30 | (0-4,669) |
|  | Women 55-74 in 2019* | 347 | (30-1,649) | 8 | (0-872) | 347 | (30-1,738) | 10 | (0-1,720) | - | - | 10 | (0-1,664) |
|  | Women 75+ in 2019 | 631 | (31-3,775) | 3 | (0-364) | 631 | (31-4,024) | 4 | (0-719) | - | - | 4 | (0-719) |
| Total |  | 15,510 |  | 14,483 |  | 16,408 |  | 15,384 |  | - |  | 11,582 |  |

PWID: people who inject drugs; MSM: men who have sex with men, a population with a high prevalence of HIV in San Francisco; TW: transgender women; SES: socioeconomic status
\*Age groups highlight the birth cohort of 'baby boomers,' which includes people born 1945-1965, who are at higher risk for undiagnosed HCV infection. [55]

**Table 2.** Summary of Estimated HCV Burden by Subpopulation, 2019. This table demonstrates the percentage of total infections borne by each subpopulation; this helps to illustrate HCV health disparities among subpopulations. For example, 73.1% of all people with anti-HCV antibodies in San Francisco in 2019 were PWID, though only an estimated 2.8% of San Francisco residents overall are PWID. Note that the rows are not exclusive (i.e., some are by risk group, some by age, and some by sex) and therefore the final three columns do not total to 100%.

| Subpopulation | # HCV seropositive |  | % with anti-HCV antibodies |  | # with untreated active HCV infection |  | % of SF population | % of citywide HCV seropositives | % of untreated active infections citywide |
| --- | --- | --- | --- | --- | --- | --- | --- | --- | --- |
|  | *pt. est.* | *(plausible range)* | *pt. est.* | *(plausible range)* | *pt. est.* | *(plausible range)* | *pt. est.* | *pt. est.* | *pt. est.* |
| PWID | **16,508** | (9,068 – 24,498) | **67.4%** | (64.6 – 70.1) | **10,468** | (4,690 – 17,628) | **2.8%** | **73.1%** | **90.4%** |
| MSM | **2,635** | (2,285 – 3,000) | **3.8%** | (3.5 – 4.1) | **119** | (0 – 423) | **7.8%** | **11.7%** | **1.0%** |
| TW (low SES) | **320** | (253 – 388) | **24.8%** | (20.3 – 29.2) | **183** | (130 – 252) | **0.1%** | **1.4%** | **1.6%** |
| Baby Boomers* | **5,982** | (3,238 – 11,430) | **3.2%** | (1.9 – 5.6) | **3,070** | (1,301 – 8,942) | **21.1%** | **26.5%** | **26.5%** |
| Men | **16,467** | (8,741 – 31,148) | **3.7%** | (2.0 – 6.8) | **7,837** | (3,179 – 21,688) | **50.8%** | **72.9%** | **67.7%** |
| Women | **5,798** | (3,019 – 12,632) | **1.3%** | (0.7 – 2.9) | **3,501** | (1,547 – 12,885) | **49.1%** | **25.7%** | **30.2%** |

PWID: people who inject drugs; MSM: men who have sex with men, a population with a high prevalence of HIV in San Francisco; TW: transgender women; SES: socioeconomic status
\*‘Baby boomers’ refer to people born 1945-1965, who were aged 55-74 in 2019, including those who are PWID, MSM, or TW, in the case of this table.

For PWID, the PSE was 24,492 people (PR: 14,037 – 34,946) with an HCV seroprevalence of 67.4% (PR: 64.6% – 70.1%); thus an estimated total of 16,508 PWID have anti-HCV antibodies (PR: 9,068 – 24,498), with an estimated 10,468 PWID having active, untreated HCV infection (PR: 4,690 – 17,628). For MSM, the PSE was 68,972 people (PR: 65,294 – 72,651) with an HCV seroprevalence of 3.8% (PR: 3.5% – 4.1%); thus an estimated total of 2,635 HCV seropositive MSM (PR: 2,285 – 3,000), with an estimated 119 MSM (PR: 0 – 423) still having untreated active infections. Of the MSM with chronic HCV infection, approximately half (52.3%) are thought to be co-infected with HIV per the 2017 MSM wave of NHBS. For low SES TW, the PSE was 1,289 (PR: 1,248 – 1,329), with an HCV seroprevalence of 24.8% (PR: 20.3% – 29.2%); thus, there was an estimated total of 320 low SES TW with anti-HCV antibodies (PR: 253 – 388), with approximately 183 TW (PR: 130 – 252) still having untreated active infections. For children ages 14 and under, the estimated total number of untreated active infections was 70 (PR: 9 – 310). Applying the ‘inflation factor’ to the Vitalant prevalence estimates for each of the general population strata resulted in a total estimate of 3,053 non-PWID, MSM, or low SES TW San Francisco residents over age 14 who are HCV seropositive (PR: 398 – 15,956), 741 of whom (PR: 35 – 16,482) had untreated active infections in 2019.

As seen in Table 2, the distribution of past and current HCV infections is highly unequal in San Francisco. People who injected drugs in the last 12 months comprised an estimated 2.8% of the San Francisco population in 2019, yet 73.1% of people with anti-HCV antibodies and 90.4% of untreated, active HCV infections in 2019 were among PWID. MSM made up an estimated 7.8% of the total population, yet 11.7% of the seropositive cases and only 1.0% of untreated active infections. Low SES TW comprised only 0.1% of the total population, but 1.4% of people with anti-HCV antibodies and 1.6% of untreated active HCV infections. Baby boomers (including those who are PWID, MSM, and TW) comprise 26.5% of both seropositive cases and untreated active infections, and only 21.1% of the total San Francisco population. Men (regardless of age or risk group) comprise 72.9% of seropositive cases and 67.7% of untreated active infections, despite being roughly half of the San Francisco population.

## Discussion

As in 2015, our updated 2019 estimate of 22,585 seropositive individuals (2.6% seroprevalence) in San Francisco is higher than the national 2018 NHANES seroprevalence estimate of 1.6% (95% CI: 1.1 – 2.4%). A higher seroprevalence is expected given San Francisco’s higher-than-average proportion of the population that are in subpopulations with increased risk of HCV infection, including PWID, MSM, and TW. Despite the high number of people with anti-HCV antibodies, however, we estimated that only 1.3% (PR: 0.6% - 4.0%) of all San Francisco residents have untreated, active HCV infection – likely a reflection of our robust efforts to diagnose infection among high-risk groups and initiate curative treatment with as many people as possible. Direct comparison of the number of untreated individuals cannot be made to 2015, as we were not able to more closely estimate the proportion of people treated and cured in that earlier estimate. However, this current analysis estimates that 48.7% (PR: 20.5 – 59.5%) of all people with anti-HCV antibodies in San Francisco no longer have active infection, far more than the ∼20-30% that would be expected to spontaneously clear [24], and therefore likely a reflection of prioritized treatment efforts since 2015.

In 2021, the SFDPH released a report of data from the Viral Hepatitis Surveillance Registry, reflecting cumulative and newly reported case reports of HCV antibodies or RNA from 2018-2019. These data showed that the newly reported cases of HCV by sex almost exactly mirror the estimated breakdown of untreated, active HCV infections in our analysis, by sex (67.1% of newly reported cases in 2018 and 67.3% of newly reported cases in 2019 were among men, compared to our estimate of 67.7% of untreated active infections among men in 2019). Not published in that report was the cumulative number of chronic HCV infections reported to SFDPH from 1 January 2007 through 31 December 2019, which was 19,538 (Amy Nishimura, personal communication). This number is 86.5% of the total number of people estimated to currently have evidence of past or present infection in San Francisco, per our results. However, while cases known to have died were removed from the registry count, there has not been a systematic cross-referencing between the HCV case registry and local or national death records; further, the current registry list includes people who moved out of San Francisco after their HCV diagnosis. Nonetheless, the relative comparability of these numbers likely indicates better-than-average HCV screening and diagnosis rates in San Francisco compared to the national average of approximately 50% nationwide [38].

Noteworthy disparities persist in San Francisco’s HCV burden, as 2019 estimates are similar to those in 2015. We found that while PWID make up only 2.8% of San Francisco’s total population, 73.1% of people with anti-HCV antibodies and 90.4% of those with untreated active infection are PWID. This is in line with a recent analysis by members of our team finding only 9.1% of young PWID in San Francisco had ever initiated HCV treatment between 2015 and 2019 [39]; however, Mirzazadeh and colleagues found that great strides have been made in HCV treatment initiation among San Francisco PWID overall, with the proportion of NHBS participants reporting having received HCV treatment increasing from only 19% in the 2015 wave to 42% in 2018 [40]. Nonetheless, the concentration of active HCV infection among PWID, combined with high rates of housing instability [41] and barriers to PWID receiving non-judgmental, high-quality medical care and prompt treatment for HCV infection [42-45], underscore an imperative to prioritize improvements in service navigation and treatment access for PWID in San Francisco and across the United States. Models of treating PWID for HCV outside of traditional clinic settings, i.e., at syringe access programs or similar venues, have already shown promise as a strategy to overcome some of these barriers and successfully cure PWID of their HCV infection [46,47].

Recent data have shown considerably higher proportions of MSM than of PWID have been treated for HCV infection, including MSM who inject drugs [39], and in this analysis only an estimated 119 MSM (PR: 0 – 423) still had untreated active HCV infections in 2019. In San Francisco, robust programs to provide HIV prevention and care services tailored to MSM may be contributing to this positive outcome, as many of these programs have incorporated HCV testing and care into their routine sexual health and/or HIV pre-exposure prophylaxis (PrEP) services. In 2019 *End Hep C SF* released a “micro-elimination plan” for HCV among people living with HIV in San Francisco (the vast majority of whom are MSM) [48], and the results from this analysis indicate that microelimination in this population may be within reach.

TW with low SES remain another San Francisco subgroup bearing a disproportionate burden of HCV infection. Despite their small absolute numbers (only 0.1% of the San Francisco population, or just over 1,000 people), low SES TW bear more than 15 times the burden of untreated HCV infection as would be expected based on their population size within San Francisco overall. Recent results from TEACH4 showed increased risk for HCV infection for TW who are older and have a history of injection drug use, and while 75% of the participants with HCV viremia reported prior knowledge of their HCV infection, only one (12%) had received treatment, which was not successfully completed [49]. Notably, almost two-thirds (63.9%) of TEACH4 participants who had anti-HCV antibodies were people of color and 35.7% identified as Black/African American [49], in a city that was 58.7% people of color and only 5.3% Black/African American in the 2020 Census [50].

Our analysis is subject to a few limitations. First, general evidence exists that there are key subgroups beyond PWID, MSM, and low SES TW that bear increased burden of HCV in San Francisco, including Blacks/African Americans, people with a history of injection drug use (but not injecting drugs in the last 12 months), people who are currently or formerly incarcerated, and people who are currently or formerly unhoused [5,51]. However, the inability to estimate prevalence of HCV seropositivity, chronic infection, or treatment for these subgroups using local data meant they were lumped into the “general population” age and sex strata. Unlike the 2015 estimate, in this analysis we did attempt to incorporate racial disparities in HCV burden through race-related reweighting of HCV prevalence in the general population strata. Nonetheless, in future updates of this analysis we hope to be able to obtain and incorporate more specific local data for these important subpopulations. Second, our triangulation of multiple data sources required numerous assumptions and extrapolations that increased estimate uncertainty. For this reason, we used ‘plausible ranges’ rather than attempting to calculate 95% confidence intervals, which imply greater statistical precision than actually exists. Further, we have made every attempt to be explicit about the areas where assumptions or extrapolations were made. Third, the overall city estimate relied heavily on local cross-sectional or longitudinal studies of key populations, each of which are subject to important selection biases. Finally, our source data (Table S3) represent timepoints across 2010-2019, rather than a single point in time (2019). Possible trends in prevalence over that time period have been ignored due to limited data availability.

Importantly, this estimate focuses on data through 2019, before the impacts of the COVID-19 pandemic were realized; some research has already demonstrated a likely negative impact on the prevention, diagnosis, and treatment of HCV in San Francisco [52] and other regions [53,54]. Regardless, as San Francisco continues to strive toward HCV elimination through *End Hep C SF*, estimates of the remaining number of people with untreated active infection are an important measure of progress. This study represents a considerable advance in methodology over the 2015 estimate and provides important information to guide planning and prioritization of limited resources to diagnose and treat HCV citywide. The methods used here could be applied by any region with local data sufficient for triangulation, to inform their own efforts to eliminate HCV.

## Data Availability

All data that can be shared publicly is already contained within the paper and/or Supporting Information files. There was no primary data collection for this study; the analysis was conducted by averaging estimates from multiple sources of information, which are either 1) explicitly stated in the paper, 2) publicly available in the source documents referenced in the paper, or 3) unpublished data from other cohorts/institutions - which was shared with permission for our use in calculations, but is not ours to publish or make publicly available on behalf of the owners of the data (the original sources are provided in S3 Table).

## Acknowledgements

We would like to thank Edward Murphy of Vitalant Research Institute, Willi McFarland of SFDPH and NHBS, Amy Nishimura of SFDPH, and Rachel McLean of the California Department of Public Health, for their assistance and insights into this analysis.

