## Supplementary material for "Hepatitis C prevalence and key population size estimate updates in San Francisco: 2015 to 2019": Table S1

### S1 Table. Literature Search Strategy.

We searched for literature from January 2017 through January 2021 in MEDLINE and Science Citation Index Expanded, to append new articles to the prior search done with the same search terms but a timeframe of 2010 through 2016. These searches resulted in 34 and 245 new articles found, respectively, with 11 unique abstracts retained for further review.

| Database | Period of Search | Search Strategy | Abstracts found | Relevant abstracts retained* |
|---|---|---|---|---|
| MEDLINE | January 2017 through January 2021 | #1 Epidemiology[MeSH] OR Incidence[MeSH] OR Prevalence[MeSH] OR Cross-Sectional Studies[MeSH] OR Cohort Studies[MeSH] OR epidemiolog*[tw] OR prevalence[tw] OR incidence[tw] OR cross-section*[tw] OR cohort*[tw]<br><br>#2 Hepatitis C[MeSH] OR hepatitis C[tw] OR hep C[tw] OR *HCV*[tw] OR anti-HCV OR HCV-RNA [tw]<br><br>#3 San Francisco [tw]<br><br>#3 AND #2 AND #1 | 34 | 7 |
| Science Citation Index Expanded (Web of Knowledge) | 2017 through 2021 [search completed on January 30, 2021] | #1 TS= (Incidence OR Prevalence OR Cross-Sectional OR Cohort OR epidemiolog*)<br><br>#2 TS = (Hepatitis C OR hep C OR HCV* OR anti-HCV OR HCV-RNA)<br><br>#3 CI = San Francisco OR PS = San Francisco<br><br>#3 AND #2 AND #1 | 245 | 10 |

***Duplicate abstracts were retained across databases; the total number of non-duplicate articles with full text found to add since last prevalence estimate in 2015 (see Table S2): 11**
