## Supplementary material for "Hepatitis C prevalence and key population size estimate updates in San Francisco: 2015 to 2019": Table S2

### S2 Table. Abstracts Retained for Further Review After Literature Search.

Eleven unique abstracts were retained for further review after following the literature search methods described in Table S1. These abstracts are listed below.

| | First Author | Title | Journal | Pub Year | Used in Analysis? |
|---|---|---|---|---|---|
| 1 | Raymond, H.F. | Estimated population size of men who have sex with men, San Francisco, 2017 | AIDS and Behavior | 2019 | yes |
| 2 | Wesson, P. | Estimating population size of transwomen in San Francisco using multiple methods, 2013. | International Journal of Transgenderism | 2017 | yes |
| 3 | Crissman, H.P. | Transgender demographics: A household probability sample of US adults, 2014. | American Journal of Public Health | 2017 | yes |
| 4 | Raymond, H.F. | Transwoman population size | American Journal of Public Health | 2017 | yes |
| 5 | Hernandez, C.J. | High hepatitis C virus seropositivity, viremia, and associated risk factors among trans women living in San Francisco, California | PloS ONE | 2021 | no, duplicate of TEACH4 dataset |
| 6 | Mirzazadeh, A. | Progress toward closing gaps in the hepatitis C virus cascade of care for people who inject drugs in San Francisco | PloS ONE | 2021 | no, duplicate of PWID NHBS 2018 dataset |
| 7 | Morris, M.D. | Treatment cascade for hepatitis C virus in young adult people who inject drugs in San Francisco: Low number treated | Drug and Alcohol Dependence | 2019 | no, duplicate of UFO dataset |
| 8 | Page, K. | HCV incidence is associated with injecting partner age and HCV serostatus mixing in young adults who inject drugs in San Francisco | PloS ONE | 2019 | No, duplicate of UFO dataset |
| 9 | Facente, S.N. | Estimated hepatitis C prevalence and key population sizes in San Francisco: A foundation for elimination | PloS ONE | 2018 | no, did not want to include estimates calculated using overlapping data in 2015 |
| 10 | Schackman, B.R. | Cost-effectiveness of hepatitis C screening and treatment linkage intervention in U.S. methadone maintenance treatment programs | Drug and Alcohol Dependence | 2018 | no, duplicate of pre-existing estimate from Perlman, *et al.* |
| 11 | Morris, M.D. | Geographic differences in temporal incidence trends of hepatitis C virus infection among people who inject drugs: The InC3 collaboration | Clinical Infectious Diseases | 2017 | No, duplicate of UFO dataset |
