## Supplementary material for "Hepatitis C prevalence and key population size estimate updates in San Francisco: 2015 to 2019": Table S3

### S3 Table. Summary of Data Sources.

The table below summarizes the main sources of data for each population size and HCV prevalence estimate contained in this analysis.

<table>
<tr><th>Subgroup</th><th>Estimate</th><th>Data source</th></tr>
<tr><td rowspan="8">PWID</td><td>PSE</td><td>Chen YH, McFarland W, Raymond HF. Estimated Number of People Who Inject Drugs in San Francisco, 2005, 2009, and 2012. AIDS and behavior. 2016;20(12):2914-21.</td></tr>
<tr><td rowspan="3">anti-HCV antibody seroprevalence</td><td>National HIV Behavioral Surveillance, San Francisco, 2018 (unpublished; available via E.W.)</td></tr>
<tr><td>UFO Study, San Francisco, 2016 (unpublished; available via M.D.M.)</td></tr>
<tr><td>Perlman DC, Jordan AE, McKnight C, Young C, Delucchi KL, Sorensen JL, et al. Viral hepatitis among drug users in methadone maintenance: associated factors, vaccination outcomes, and interventions. Journal of addictive diseases. 2014;33(4):322-31.</td></tr>
<tr><td rowspan="2">HCV RNA and acute prevalence</td><td>National HIV Behavioral Surveillance, San Francisco, 2018 (unpublished; available via E.W.) [RNA prevalence only]</td></tr>
<tr><td>UFO Study, San Francisco, 2016 (unpublished; available via M.D.M.)</td></tr>
<tr><td rowspan="2">proportion treated</td><td>National HIV Behavioral Surveillance, San Francisco, 2018 (unpublished; available via E.W.)</td></tr>
<tr><td>UFO Study, San Francisco, 2016 (unpublished; available via M.D.M.)</td></tr>
<tr><td rowspan="9">MSM</td><td rowspan="3">PSE</td><td>Grey JA, Bernstein KT, Sullivan PS, Purcell DW, Chesson HW, Gift TL, et al. Estimating the Population Sizes of Men Who Have Sex With Men in US States and Counties Using Data From the American Community Survey. JMIR Public Health Surveill. 2016;2(1):e14.</td></tr>
<tr><td>Hughes A, Chen YH, Scheer S, Raymond HF. A novel modeling approach for estimating patterns of migration into and out of San Francisco by HIV status and race among men who have sex with men. Journal of Urban Health. 2017;94(3):350-363.</td></tr>
<tr><td>Raymond HF, McFarland W, Wesson P. Estimated Population Size of Men Who Have Sex with Men, San Francisco, 2017. AIDS and behavior. 2019;23(6):1576-9.</td></tr>
<tr><td rowspan="3">anti-HCV antibody seroprevalence</td><td>National HIV Behavioral Surveillance, San Francisco, 2017 (unpublished; available via E.W.)</td></tr>
<tr><td>Electronic medical record data from Strut sexual health clinic, San Francisco AIDS Foundation, 2017-2018 (unpublished; available via J.H.)</td></tr>
<tr><td>Community-based and clinic-based HCV testing program data, San Francisco Department of Public Health, 2019 (unpublished; available via R.G.)</td></tr>
<tr><td>HCV RNA prevalence</td><td>Community-based and clinic-based HCV testing program data, San Francisco Department of Public Health, 2019 (unpublished; available via R.G.)</td></tr>
<tr><td rowspan="2">proportion treated</td><td>National HIV Behavioral Surveillance, San Francisco, 2017 (unpublished; available via E.W.)</td></tr>
<tr><td>Electronic medical record data from Strut sexual health clinic, San Francisco AIDS Foundation, 2017-2018 (unpublished; available via J.H.)</td></tr>
</table>

| | | |
|---|---|---|
| TW | PSE | Wesson P, Oabazard R, Wilson EC, McFarland W, Raymond HF. Estimating population size of transwomen in San Francisco using multiple methods, 2013. International Journal of Transgenderism. 2017. |
| | | Crissman HP, Berger MB, Graham LF, Dalton VK. Transgender Demographics: A Household Probability Sample of US Adults, 2014. American journal of public health. 2017;107(2):213-5. |
| | | Raymond HF, Wilson EC, McFarland W. Transwoman Population Size. American journal of public health. 2017;107(9):e12. |
| | anti-HCV antibody seroprevalence | TEACH3 cohort, San Francisco, 2017 (unpublished; available via E.W.) |
| | | TEACH4 cohort, San Francisco, 2019 (unpublished; available via E.W.) |
| | HCV RNA prevalence | TEACH3 cohort, San Francisco, 2017 (unpublished; available via E.W.) |
| | | TEACH4 cohort, San Francisco, 2019 (unpublished; available via E.W.) |
| | proportion treated | TEACH3 cohort, San Francisco, 2017 (unpublished; available via E.W.) |
| | | TEACH4 cohort, San Francisco, 2019 (unpublished; available via E.W.) |
| General Population | PSE | United States Census Bureau / American Fact Finder. Age and Sex: 2019 American Community Survey 1-Year Estimates. U.S. Census Bureau's American Community Survey Office, 2019. |
| | HCV prevalence | Data for first-time allogeneic blood donors at the Vitalant Systems Research Institute (BSRI) from 2010-2019 who were San Francisco residents at the time of donation (unpublished; available via R.B and Z.K.) |
| | | Steele WR, Dodd RY, Notari EP, Xu M, Nelson D, Kessler DA, et al. Prevalence of human immunodeficiency virus, hepatitis B virus, and hepatitis C virus in United States blood donations, 2015 to 2019: The Transfusion-Transmissible Infections Monitoring System (TTIMS). Transfusion. 2020;60(10):2327-39. |
| | | National Center for Health Statistics, Centers for Disease Control and Prevention (CDC). National Health and Nutrition Examination Survey Data (NHANES) 2018. Hyattsville, MD. |
